# From National Data to Local Evaluation: Benchmarking Machine Learning Models for Perioperative Risk Prediction

**DOI:** 10.64898/2026.09.02.26361824

**Authors:** Zihan Ding, Grace Han, Tengfei Ma, Fusheng Wang, Janos Hajagos, Tahsin Kurc, Anisha R. Kumar

**Affiliations:** Department of Computer Science, Stony Brook University, Stony Brook, New York; Department of Biomedical Informatics, Stony Brook University, Stony Brook, New York; Renaissance School of Medicine, Stony Brook University, Stony Brook, New York; Division of Otolaryngology, Department of Surgery, Renaissance School of Medicine, Stony Brook University, Stony Brook, New York

## Abstract

Whether newer modeling approaches improve perioperative risk prediction is unclear. Holding 73 prespecified preoperative variables fixed — 69 observed and used as tabular inputs, all 73 serialized to text — we benchmarked six tabular model families and three pretrained clinical text-encoder embedding classifiers for four 30-day outcomes, training on 4,995,670 ACS NSQIP cases (2018–2022) and testing temporally on 963,565 cases (2024). The FT-Transformer led or tied on AUROC for all four outcomes (0.755–0.955), exceeding logistic regression by only 0.009–0.030; the three encoders fell 0.005–0.009 short of the best tabular model and spanned 0.003 or less. On identical cases the ACS NSQIP Surgical Risk Calculator matched the best model for mortality but trailed for any complication. Rankings changed under AUPRC and did not hold in a 3,490-case single-institution deployment check, not independent of the PUF. Once the predictor set is fixed, architecture and encoder choice move discrimination little.

## Introduction

Perioperative risk prediction informs surgical decision-making, informed consent, and resource allocation across the preoperative, intraoperative, and postoperative periods. Current perioperative risk tools, including the American College of Surgeons (ACS) National Surgical Quality Improvement Program (NSQIP) Surgical Risk Calculator, rely exclusively on structured variables such as age, comorbidities, and procedure type^1^. The calculator’s discrimination and calibration have been examined and refined on the national sample it was built from^2, 3^, but its underlying architecture — a set of hierarchical regression models — has not been systematically benchmarked against newer modeling paradigms, including deep learning, using the same cohort and outcome definitions.

Perioperative outcomes are not uniformly distributed across sociodemographic groups. A recent multi-institutional analysis of over 7.5 million ACS NSQIP patients found that Black and American Indian/Alaska Native patients experienced higher rates of reoperation, wound healing complications, and other adverse postoperative outcomes compared with White patients, while Asian patients generally experienced comparable or better outcomes^4^. Because demographic variables such as race, sex, and age are included among this study’s predictors, any disparities reflected in the training data have the potential to be learned and reproduced by the benchmarked models, as has been documented for deployed health care algorithms^5^; this motivates future evaluation of model performance across sociodemographic subgroups (see Limitations section).

Prior work has directly compared modeling architecture on structured perioperative data, holding predictors constant: deep learning has been benchmarked against random forest and XGBoost for nine postoperative complications using shared preoperative, intraoperative, and perioperative variables^6^, and XGBoost, light-gradient boosting, multilayer perceptron, logistic regression, and balanced random forest have been compared for postoperative pulmonary edema across a five-hospital cohort with external validation^7^. Related single-center work has shown that models fit to routinely collected preoperative structured data can predict postoperative mortality with high discrimination, whether through deep networks^8^ or automated tree-based pipelines^9^. These studies establish that architecture-isolated comparisons on structured data are feasible and clinically informative, but none includes a classifier built on embeddings from a pretrained clinical language model alongside the conventional architectures.

Beyond the perioperative setting, the broader tabular-data literature offers conflicting guidance on whether deep learning architectures outperform gradient-boosted trees. A large-scale benchmark across 45 tabular datasets found that tree-based models remain state-of-the-art on medium-sized data even without accounting for their faster training^10^, and a comparison across 11 datasets reached a similar conclusion^11^, while a separate benchmark introducing the FT-Transformer architecture found it competitive with, though not uniformly superior to, gradient-boosted decision trees across a diverse set of tabular tasks^12^. This unresolved comparison motivates evaluating both tree-based and transformer-based architectures directly on perioperative outcome prediction rather than assuming either paradigm’s superiority.

Representation, not only architecture, has been a persistent theme in EHR prediction. Mapping the raw record into a standard sequential format and letting a deep model consume it directly, without hand-curated features or site-specific harmonization, predicted inpatient mortality, readmission, length of stay and discharge diagnoses across two academic medical centers^13^, establishing that how the record is presented to the model is itself a design choice with measurable consequences.

A separate line of work asks whether structured records are better represented as text than as a feature vector. Serializing each tabular row into a natural language sentence and handing it to a language model was introduced as a few-shot method for tabular classification^14^ and has since been applied to medical records directly: extracting frozen embeddings from templated text representations of structured electronic health record (EHR) values and training a conventional classifier on those embeddings recovered much, though not all, of the discrimination available from the raw numerical features themselves^15^. A more recent evaluation converting EHR codes to plain-text descriptions and embedding them with general-purpose and clinical encoders found the resulting representations competitive with a dedicated EHR foundation model across 15 tasks, while also finding that gradient boosting on the original structured features remained strong where those features were complete^16^. This is precisely the design evaluated here, and it has not been tested on perioperative outcomes.

The encoders used for that purpose should be distinguished from generative large language models (LLMs) prompted to produce a prediction. A health system-scale language model trained on clinical notes performed comparably to task-specific models across multiple operational and clinical prediction tasks^17^, whereas a benchmark comparing general-purpose generative LLMs (GPT-3.5, GPT-4) against locally trained machine learning models on EHR-based prediction tasks found that traditional machine learning substantially outperformed non-fine-tuned LLMs in both discrimination and calibration^18^. Together these findings indicate that language-model performance on clinical prediction depends heavily on how the model is used — prompted, fine-tuned, or read as a frozen feature extractor — rather than being uniformly competitive with conventional approaches, and they motivate evaluating the frozen-embedding configuration on perioperative outcomes under the same predictors and evaluation framework as the tabular models.

Building on the architecture-isolated comparisons above, this study extends that comparison to include classifiers trained on frozen embeddings from pretrained clinical text encoders, using structured data alone: no study to our knowledge has directly compared traditional machine learning, conventional deep architectures, and text-encoder embeddings of serialized structured records within the same predictors and outcome definitions. This comparison establishes a comparative baseline across modeling paradigms, so that observed differences reflect modeling approach rather than differences in cohort composition or outcome definition.

This study benchmarks six tabular model families spanning traditional machine learning and deep learning, together with three pretrained clinical text-encoder embedding classifiers, on structured perioperative data for four outcomes: 30-day readmission, return to the operating room (OR) within 30 days, 30-day mortality, and any complication within 30 days, using the national ACS NSQIP PUF cohort. Because this cohort permits no reliable patient-level identification, training and test sets are separated along the time dimension rather than split randomly, which also matches the temporal shift a deployed model faces^19^ (see Methods). This comparison establishes baseline performance for each modeling paradigm on structured data alone, before free-text clinical documentation and imaging can be incorporated in future work. Isolating the contribution of modeling approach from that of these additional data modalities clarifies which architectural choices merit further investment as the framework is extended to unstructured and imaging data.

The objective of this study is to compare the discriminative performance of traditional machine learning, conventional deep learning, and frozen clinical text-encoder embeddings for four perioperative outcomes, using the national ACS NSQIP PUF cohort.

## Methods

This study was designed and is reported in accordance with the TRIPOD+AI statement for prediction model studies using regression or machine learning methods^20^. This study was designed and is reported in accordance with the TRIPOD+AI statement for prediction model studies using regression or machine learning methods; the completed checklist is provided in Supplementary Table S1. This study was prospectively registered on the Open Science Framework prior to model training and evaluation (Registration DOI: 10.17605/OSF.IO/JVKDH).

### Study Design and Data Source

This retrospective benchmarking study compared nine modeling configurations — six tabular model families spanning traditional machine learning and deep learning, and three classifiers trained on frozen embeddings from pretrained clinical text encoders — for prediction of four 30-day postoperative outcomes. All models used the same clinician-reviewed preoperative predictor set, outcome definitions, training and test cohorts, and evaluation pipeline, allowing model performance to be compared while holding the underlying clinical information constant. Six models operated directly on structured tabular inputs, whereas the three encoder configurations represented the same structured variables as narrative text using a fixed serialization template, following the structured-to-text serialization approach used for tabular^14^ and electronic health record data^15, 16^.

The source dataset was the ACS NSQIP Participant Use File (PUF) for operative years 2018–2024, comprising 6,953,549 cases from more than 700 participating hospitals^21^. The ACS NSQIP Participant Use File is a publicly available, fully de-identified dataset distributed by the American College of Surgeons; because it contains no institution, facility, or patient-level identifiers, its use did not require IRB review or approval.

The PUF contains a systematically sampled subset of operations performed at participating sites rather than all surgical procedures, and records the operative year but not the month or exact date of surgery. Prediction was performed at the operative-case level rather than the patient level. This is consistent with the unit of prediction used by the ACS NSQIP Surgical Risk Calculator and is also required by the absence of patient-level identifiers in PUF.

### Cohort Definition

The development cohort included 4,995,670 PUF cases from 2018–2022, and temporal evaluation was performed on 963,565 cases from 2024. The 994,314 cases from 2023 were excluded as a prespecified one-year temporal gap, used to increase separation between the development and evaluation periods and to reduce potential temporal proximity or overlap between them. Because PUF records the operative year but neither exact dates nor a patient identifier, the gap cannot establish that the two cohorts contain disjoint patients, and it is not claimed to prevent patient-level leakage; it widens the interval over which case mix, coding and practice would have to remain stable for the evaluation to be optimistic. No additional eligibility criteria were applied, and no cases were excluded on clinical grounds.

Performance was additionally assessed in a single-institution cohort of 3,490 cases performed at one academic medical center in 2024–2025 and harmonized to the PUF variable definitions and coding scheme. Two further cases were excluded during harmonization: one carried a CPT code not observed in any PUF year and therefore had no ACS work relative value unit, and one had missing race. This cohort is not independent of the PUF sample. The contributing hospital participates in ACS NSQIP, and because the PUF is a systematic sample drawn from participating sites, an unknown subset of these cases may also be represented in the PUF evaluation year; PUF carries no institution identifier, so the overlap can be neither measured nor removed. This analysis is therefore reported as a single-institution deployment check rather than as external validation, and the two evaluation cohorts should not be read as independent evidence. Its case mix also differs from the national sample, with a higher proportion of female patients (69.2% versus 57.9%) and slightly younger patients (Table 1).

**Table 1.**
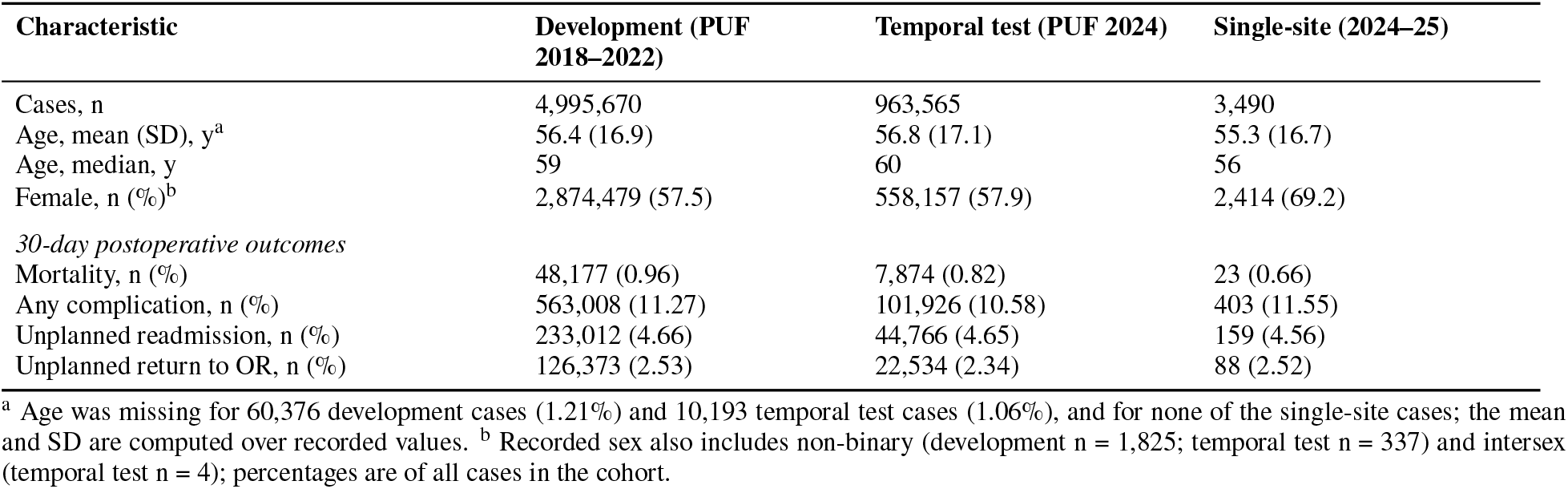
Baseline characteristics and outcome frequencies for the development, temporal test, and single-institution cohorts.

No prospective sample size calculation was performed. Study size was instead determined by all eligible cases available in the ACS NSQIP PUF across the specified development (2018–2022) and evaluation (2024) periods, an externally collected, multi-institutional dataset assembled independent of this study. This substantial sample size (4,995,670 development cases; 963,565 test cases) was considered sufficient to detect clinically meaningful differences in discrimination between architectures with narrow confidence intervals.

### Outcomes

Four binary outcomes were evaluated over a 30-day postoperative window: (1) unplanned readmission, defined as any of UNPLANNEDREADMISSION1 through UNPLANNEDREADMISSION5 recorded as “Yes” with a corresponding READMPODAYS value of 30 days or fewer; (2) unplanned return to the operating room, defined as RETURNOR recorded as “Yes” with RETORPODAYS of 30 days or fewer; (3) mortality, defined by DOpertoD (days from operation to death) containing a value other than the −99 sentinel, which NSQIP uses for cases without death within 30 days; and (4) any complication, defined as the occurrence of at least one of 18 NSQIP postoperative occurrence variables. Readmissions and returns to the operating room were counted regardless of whether they were judged related to the principal procedure.

The complication composite included superficial incisional (SUPINFEC), deep incisional (WNDINFD), and organ/space (ORGSPCSSI) surgical site infection; wound dehiscence (DEHIS); pneumonia (OUPNEUMO); unplanned intubation (REINTUB); pulmonary embolism (PULEMBOL); ventilator dependence beyond 48 hours (FAILWEAN); progressive renal insufficiency (RENAINSF); acute renal failure (OPRENAFL); urinary tract infection (URNINFEC); stroke (CNSCVA); cardiac arrest requiring cardiopulmonary resuscitation (CDARREST); myocardial infarction (CDMI); bleeding requiring transfusion (OTHBLEED); deep venous thrombosis (OTHDVT); sepsis (OTHSYSEP); and septic shock (OTHSESHOCK). Occurrences flagged by NSQIP as present at the time of surgery were retained. Outcomes were recorded prospectively by NSQIP abstractors independent of this study’s model development, so no separate blinding procedure was required.

All variables used to derive study outcomes, including day-offset fields, readmission and reoperation detail fields, and postoperative occurrence variables, were excluded from model input to prevent outcome leakage. ACS-supplied MORTPROB and MORBPROB estimates were also excluded because they represent predictions from an existing risk model rather than primary preoperative predictors.

### Predictors and Data Preprocessing

Predictors were restricted to variables available before the start of surgery. We prespecified 73 preoperative PUF variables, finalized before model development and fixed thereafter, spanning demographics and anthropometry, operative context, comorbidities and clinical status, and preoperative laboratory measurements (Table 2). This predictor set was selected by a single clinician and co-author (A.R.K.) based on clinical relevance to perioperative risk, rather than derived from a previously published predictor set or risk model. Sixty-nine of the 73 had observed values in the training years and were used as tabular inputs; all 73 field slots were serialized for the embedding models, with the remaining four — DPRHEMOGLOBIN, DPRHEMO_A1C, PRPT and DPRPT — uniformly represented as undefined. Those four therefore contributed no information to any model in either arm, and the two arms saw the same clinical content. This 69-versus-73 accounting is used consistently throughout: 69 instantiated tabular inputs, 73 serialized field slots. No intraoperative or postoperative variables were used.

**Table 2.** Composition of the clinician-reviewed preoperative predictor set. Sixty-nine of the 73 had observed values in the training years and were used as tabular inputs; all 73 field slots were serialized for the embedding models, the last four uniformly as undefined.

| Predictor domain | n | Description |
| --- | --- | --- |
| Demographics and anthropometry | 6 | Age, sex, race/ethnicity, height, weight |
| Operative context | 11 | Procedure, specialty, setting, urgency, transfer status |
| Cardiopulmonary and renal comorbidity | 8 | Pulmonary, cardiac, renal, and respiratory status |
| Systemic, oncologic, and infectious status | 12 | Metabolic, oncologic, immune, infectious, and bleeding conditions |
| Functional and geriatric status | 4 | Functional status, dementia, falls, home support |
| Preoperative laboratory variables | 28 | Laboratory values and timing before surgery |
| Prespecified variables with no observed training values | 4 | No observed value in training years |
| **Clinician-reviewed predictor set** | **73** |  |

Variable availability and coding changed across NSQIP years and between the national and single-institution cohorts. Table 3 gives each predictor’s availability in the three cohorts alongside the harmonization applied. Three classes of difference arose: variables discontinued or introduced during the study period, variables requiring recoding across PUF eras, and one variable measured differently in the two cohorts — preoperative haemoglobin, which PUF derives from haematocrit whereas the single-institution records measure it directly. The prespecified predictor set was retained despite these differences, with unavailable values handled through the fitted preprocessing pipeline rather than by dropping predictors.

**Table 3.**
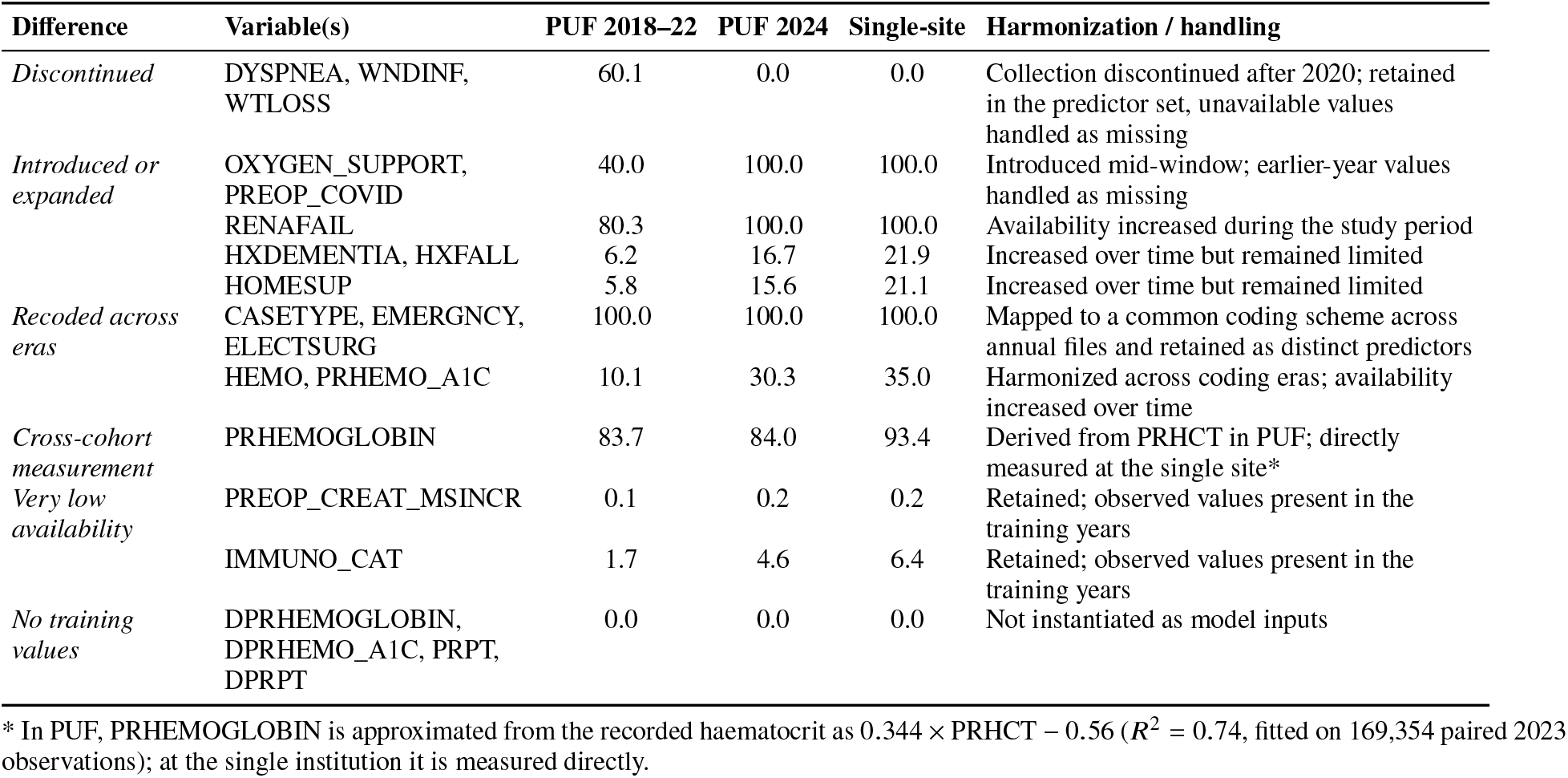
Temporal and cross-cohort differences in predictor availability and harmonization. Values are the percentage of cases with a recorded value in each cohort.

PUF sentinel values of −99 were converted to missing for numeric variables. All preprocessing parameters, including imputation values, standardization constants, category levels, and vocabularies, were fitted only on the 95% fit split of the development cohort and then applied without refitting to the validation split and the 2024 temporal test cohort. Logistic regression used median imputation and standardization for numeric predictors, with one-hot encoding for categorical variables with 30 or fewer levels and frequency encoding for higher-cardinality variables. Random forest used median-imputed numeric predictors and integer-coded categorical variables. XGBoost and LightGBM retained numeric missing values and used fixed categorical levels. The multilayer perceptron and FT-Transformer used median-imputed, standardized numeric predictors and categorical vocabularies capped at 1,000 levels per variable, with a reserved index for unseen categories. For the embedding classifiers, each patient’s record was serialized into narrative text using a single fixed template that names all 73 prespecified field slots in the data file’s column order and pairs each with the recorded value; the template wording was transcribed from the ACS NSQIP data dictionary. Missing values fill their slot with the literal token undefined rather than being imputed or omitted, so the passage has the same fields in the same order for every patient in every cohort. The four field slots with no observed training values were written as undefined in all cohorts, matching the information available to the tabular models, which do not instantiate them.

The serialized passage exceeds the 512-position limit shared by all three encoders (1,029 to 1,073 tokens, depending on the tokenizer), so the 73 field slots were partitioned into three fixed contiguous groups, each rendered as its own standalone passage of at most 512 tokens. The partition is identical for every patient, cohort and encoder, and was derived from worst-case per-field token cost measured across the three tokenizers on 4,000 training records; no field is split across a boundary and none is dropped. Zero truncation was verified on all three cohorts before any embedding was generated. Each passage was passed through the frozen encoder, the three resulting vectors were L2-normalized and averaged, and the mean was L2-normalized again to give one representation per patient.

### Models and Training

The six tabular families were L2-regularized logistic regression, random forest^22^, XGBoost^23^, LightGBM^24^, a multi-layer perceptron, and the FT-Transformer^12^; together they span the linear, tree-ensemble and deep architectures whose relative merits on tabular data remain disputed^10, 11^.

Hyperparameters were prespecified rather than searched. A single configuration per model family — fixed before any model was fit, carried over unchanged from earlier experiments on this dataset, and identical across the four outcomes — was used for every run; the values are given under Model Parameters below. No grid, random, or Bayesian search was performed, and no hyperparameter value was chosen on the basis of an observed performance number. The 5% validation split therefore served a single circumscribed purpose: early stopping for the families that support it. For XGBoost the number of boosting rounds was chosen by early stopping on validation AUPRC (stop after 50 rounds without improvement; maximum 800). For LightGBM the criterion was validation average precision, with early stopping restricted to that one metric (50 rounds; maximum 1,500). For the MLP and the FT-Transformer, training stopped when validation AUROC had not improved for five consecutive epochs (maximum 30 epochs), and the parameters from the best epoch were restored. Logistic regression and random forest do not early-stop and never read the validation rows; those rows were nonetheless excluded from their fit, so all six families were trained on identical rows and the between-model comparison reflects the model rather than the data it saw. No family was discarded on validation performance: all six were carried forward and are reported.

Class imbalance was addressed by cost-sensitive re-weighting rather than by resampling; no rows were up-sampled, down-sampled, or synthesised. Within each outcome, positive cases were weighted in inverse proportion to class frequency in the fit set, giving an effective positive-to-negative weight ratio equal to that outcome’s negative-to-positive count ratio (mortality 102.7, unplanned reoperation 38.5, readmission 20.4, any complication 7.9). This was implemented as `class_weight=“balanced”` (logistic regression), `class_weight=“balanced_subsample”` (random forest), `scale_pos_weight` (XGBoost, LightGBM), and a positive-class weight in the binary cross-entropy loss (MLP, FT-Transformer). Re-weighting is not a neutral transformation: it changes what each model fits and can therefore change the ranking it produces, so AUROC and AUPRC are not invariant to it, and it distorts the output scale so that the resulting scores are not calibrated absolute risks^25^. The weighted-loss outputs are accordingly interpreted as rankings only. AUROC and AUPRC are threshold-independent discrimination metrics and do not require calibrated probability estimates, which is why they are the primary metrics here. No probability calibration was applied; calibration measures (Brier score, log loss, calibration slope, ECE) are consequently neither computed nor reported^26^, and every operating point is rank-based rather than a fixed absolute-risk cut-off. Because the same weighting scheme was applied identically to all nine configurations within each outcome, it does not favour any one of them in the comparison.

Three pretrained text encoders were used as frozen feature extractors. All three are BERT-family bidirectional encoders^27^ rather than generative large language models, and we refer to this arm as the text-encoder embedding classifiers throughout. Each was obtained from HuggingFace and pinned to a specific commit, since authors update weights in place under an unchanged name. (i) `abhinand/MedEmbed-large-v0.1` (commit 963121b), a 335M-parameter retrieval embedding model fine-tuned from BAAI/bge-large-en-v1.5 on medical text; it has no peer-reviewed description and is cited as a model resource by its model card and commit^28^. (ii) `medicalai/ClinicalBERT` (commit f7c7f65), a 110M-parameter masked-language checkpoint whose card reports pretraining on a multicentre corpus of 1.2 billion words and fine-tuning on approximately 3 million electronic health records; we could not identify a peer-reviewed publication describing this checkpoint, so it too is cited as a model resource by card and commit^29^, and it should not be conflated with other checkpoints distributed under the name ClinicalBERT. (iii) `emilyalsentzer/Bio_ClinicalBERT` (commit d5892b3), a 110M-parameter checkpoint initialized from BioBERT^30^ and further pretrained on the clinical notes of MIMIC-III^31^, a single-centre intensive care database^32^. No encoder weight was updated at any point. Pooling followed each model card rather than being selected on performance: the CLS position for MedEmbed-large-v0.1, which is trained as an embedding model, and the attention-masked mean of the final hidden states for the two masked-language checkpoints, whose CLS token was never trained as a sentence representation.

The downstream classifier is the multilayer perceptron described above, used without modification: the same 512–256–128 trunk, the same dropout, the same optimizer, batch size, epoch cap and early stopping on validation AUROC, and the same outcome-specific positive-class weighting. Embedding dimensions were standardized using means and standard deviations fitted on the fit split alone, the same treatment the tabular models’ numeric predictors receive. The four outcomes, the fit/validation split, the 2024 temporal test cohort and the single-institution cohort are identical to those used by the six tabular families, so the comparison between the two isolates the input representation and nothing else. Every model was evaluated on the complete 2024 test cohort; no sampling was applied.

### Model Parameters

The prespecified configuration of each family was as follows. Logistic regression used the saga solver with an L2 penalty, C = 1.0, at most 200 iterations and a tolerance of 10^−3^. The random forest grew 300 trees to a maximum depth of 20, requiring at least 50 samples per leaf and considering 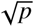 features at each split. XGBoost used a learning rate of 0.05, maximum depth 8, row and column subsampling of 0.8, a minimum child weight of 5, λ = 1.0 and the hist tree method, with AUPRC as the evaluation metric. LightGBM used a learning rate of 0.05, 127 leaves, unrestricted depth, row subsampling of 0.8 applied at every iteration, column subsampling of 0.8, at least 100 samples per leaf and λ = 1.0.

Restricting LightGBM’s early stopping to average precision is not cosmetic. Because every model is trained with class re-weighting, validation binary log loss is best at the first iteration and degrades thereafter; left at its default, LightGBM tracked log loss alongside average precision and stopped at a single tree.

The multilayer perceptron had hidden layers of 512, 256 and 128 units, with hidden dropout 0.3 and embedding dropout 0.1. The FT-Transformer used a token dimension of 64, three blocks, eight attention heads, attention dropout 0.2, feed-forward dropout 0.1, no residual dropout, and a feed-forward factor of 4/3. The two neural families shared a training configuration: batch size 8,192 — 2,048 for the FT-Transformer — a learning rate of 10^−3^ and weight decay 10^−5^; their epoch budget and early-stopping rule are given above, as are the class weights applied to every family.

The three text-encoder embedding classifiers introduce no further hyperparameters: the encoders are frozen, and the downstream classifier is the multilayer perceptron described above, used unchanged.

### Statistical Analysis

Model performance was evaluated using the area under the receiver operating characteristic curve (AUROC) and the area under the precision-recall curve (AUPRC) for each of the four outcomes. AUPRC is reported alongside AUROC because all four outcomes are heavily imbalanced, a setting in which the ROC curve alone can present an over-optimistic picture^33^. To place these values against the tool currently in clinical use, ACS NSQIP’s own precomputed risk estimates were scored on the identical 2024 test cases: MORTPROB for mortality and MORBPROB for any complication. Both fields were populated for all 963,565 cases, so this reference is computed on exactly the rows used for every model. These estimates were excluded from model input throughout and were not refitted; the calculator publishes no analogous estimate for unplanned return to OR or readmission, so the comparison covers two of the four outcomes. Ninety-five percent confidence intervals were estimated by nonparametric bootstrap resampling of each evaluation cohort with replacement for 1,000 replicates^34^. Performance was assessed in the 2024 PUF temporal test cohort using frozen preprocessing and model parameters, without refitting or recalibration, which is the temporal form of model validation^35, 36^; because the single-institution cohort is not independent of the PUF sample, neither cohort constitutes external validation in that sense. No specific approach to address model fairness across sociodemographic subgroups (e.g., stratified performance evaluation) was undertaken in this benchmarking study.

## Results

### Tabular Model Performance

In the 2024 PUF temporal test cohort, the FT-Transformer achieved the highest AUROC for mortality (0.955), any complication (0.822), and readmission (0.755), and tied with the MLP for unplanned return to OR (0.777) (Table 4, AUROC rows). Logistic regression had the lowest AUROC across all four outcomes; the absolute difference between FT-Transformer and logistic regression ranged from 0.009 for mortality to 0.030 for unplanned reoperation. Differences among the nonlinear models were smaller, particularly for any complication, for which XGBoost, LightGBM, MLP, and FT-Transformer achieved AUROCs of 0.821–0.822. Rankings differed under AUPRC (Table 4, AUPRC rows): LightGBM had the highest mortality AUPRC (0.302), XGBoost and LightGBM tied for any complication (0.461), MLP and FT-Transformer tied for unplanned return to OR (0.095), and MLP achieved the highest readmission AUPRC (0.135).

**Table 4.**
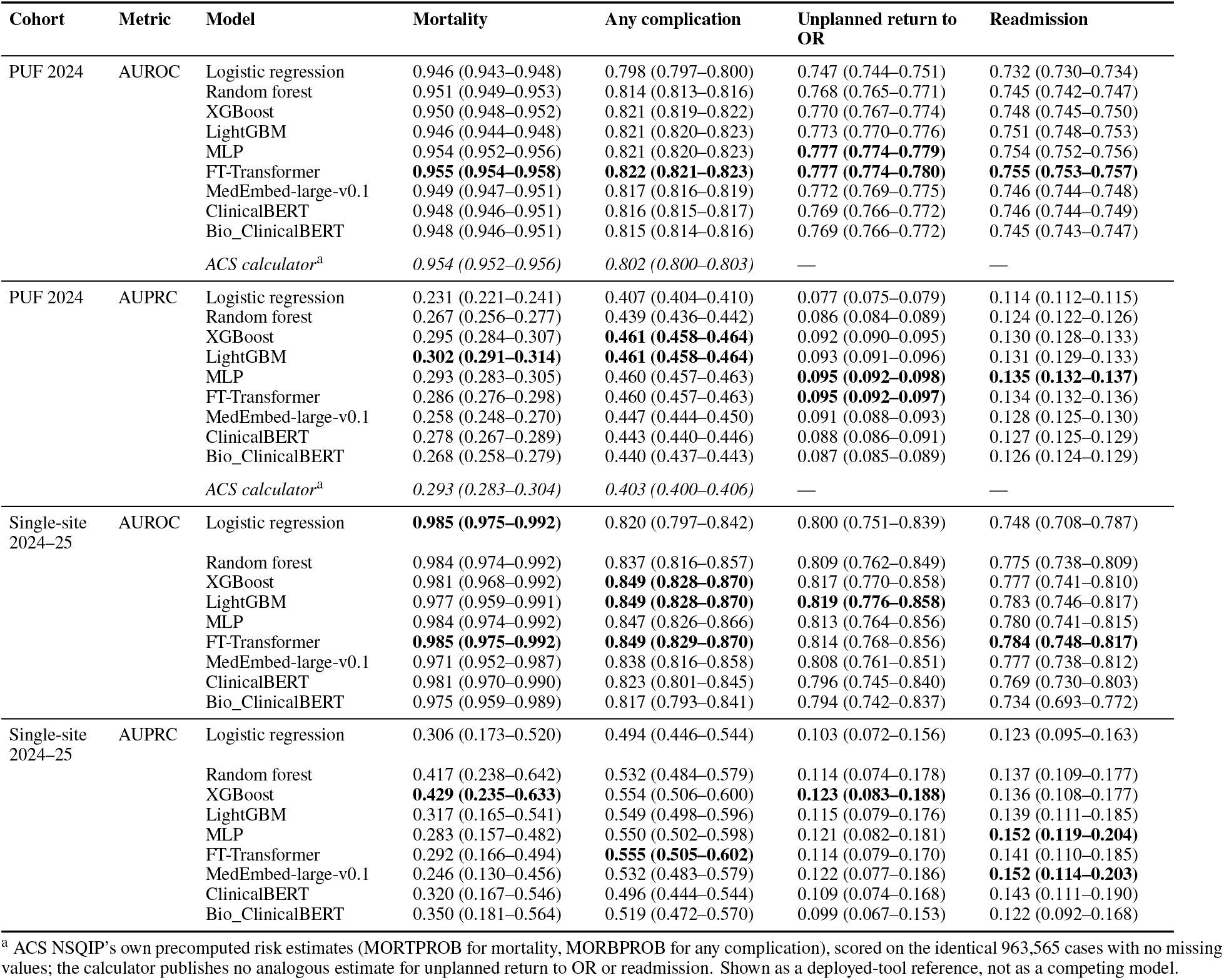
Discrimination of all nine modelling approaches in the 2024 PUF temporal test cohort and the single-institution cohort. The same frozen models and preprocessing were applied to both cohorts without refitting or recalibration. Values are AUROC or AUPRC with 95% confidence intervals from 1,000 nonparametric bootstrap resamples, computed by one implementation for all nine models. Bold indicates the highest point estimate within each outcome, cohort and metric, across the nine models; ties are bolded, and the ACS reference row is excluded from bolding. The single-institution cohort is not independent of the PUF sample (see Methods).

Measured against the deployed ACS NSQIP risk calculator on the identical cases, the picture differed sharply by outcome. For mortality the calculator’s own estimates reached an AUROC of 0.954 (0.952–0.956), within 0.001 of the best of the nine approaches (FT-Transformer, 0.955), and an AUPRC of 0.293 (0.283–0.304) against LightGBM’s 0.302 (0.291–0.314). For any complication the trained models were clearly ahead: 0.822 (0.821–0.823) versus 0.802 (0.800–0.803) by AUROC and 0.461 (0.458–0.464) versus 0.403 (0.400–0.406) by AUPRC, with non-overlapping confidence intervals on both metrics.

**Figure 1.**
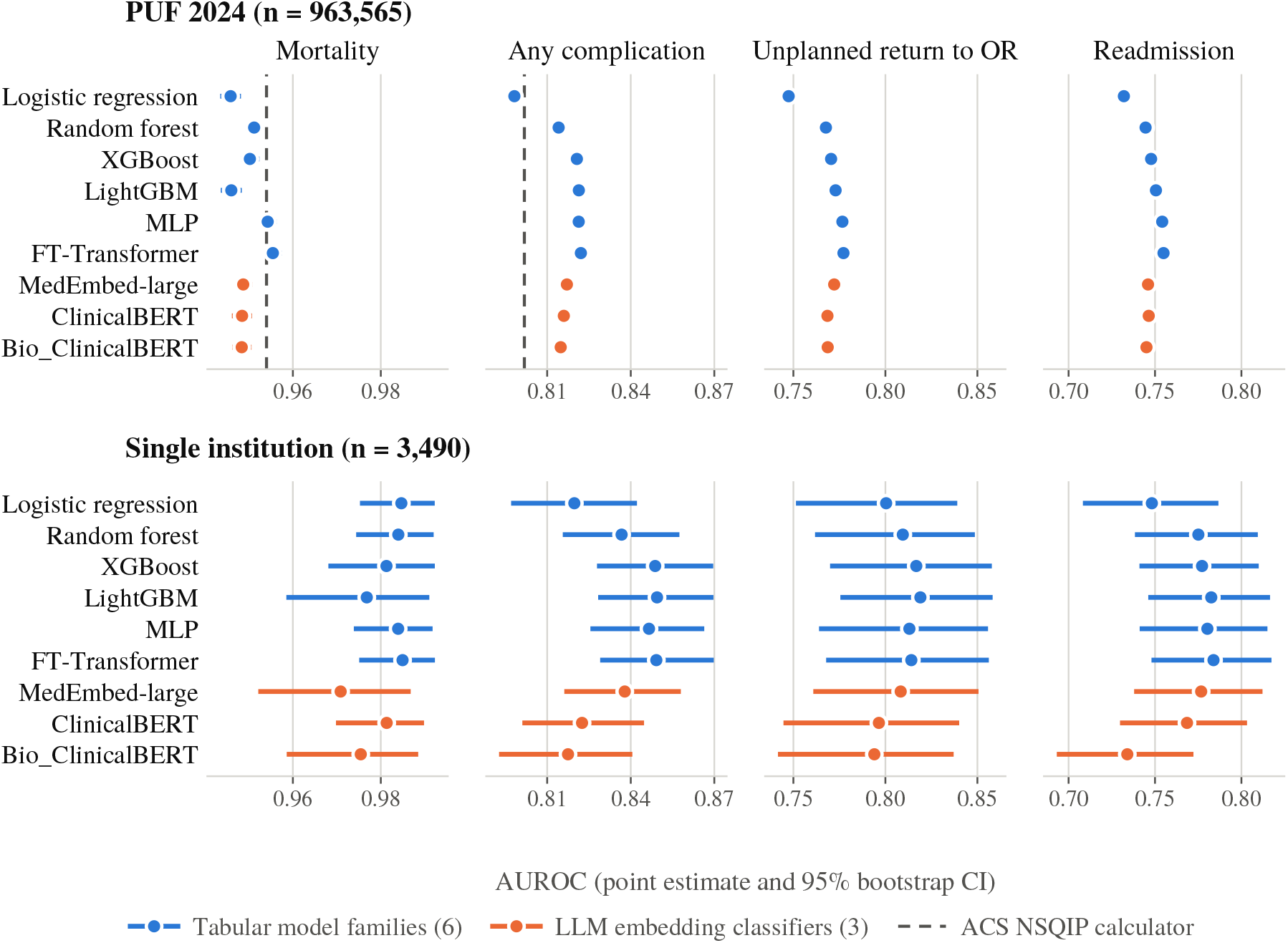
AUROC of all nine models in the 2024 PUF temporal test cohort (top) and the single-institution cohort (bottom), with 95% bootstrap confidence intervals. Within each outcome the two rows share one horizontal scale; scales differ between outcomes. The dashed rule is the ACS NSQIP calculator scored on the identical cases.

### Single-Institution Cohort

The same frozen models and preprocessing were applied without refitting to the 3,490-case single-institution cohort (Table 4). AUROC point estimates were higher there than in the national test cohort for every outcome. This is consistent with a difference in case mix rather than with better generalization, and the cohort is not independent of the PUF sample (see Methods), so it does not test transfer to an unseen institution. Confidence intervals were substantially wider and the ordering observed nationally did not hold: logistic regression and the FT-Transformer tied for the highest mortality AUROC (0.985), XGBoost, LightGBM and the FT-Transformer tied for any complication (0.849), LightGBM led for unplanned return to OR (0.819), and the FT-Transformer led for readmission (0.784). Under AUPRC, XGBoost led for mortality (0.429) and unplanned return to OR (0.123), the FT-Transformer for any complication (0.555), and the MLP for readmission (0.152). Intervals overlapped across all six families for every outcome; for mortality, with 23 events, all six point estimates lay between 0.977 and 0.985 and the AUPRC intervals were as wide as 0.157–0.642. These results therefore cannot rank architectures, and are reported to show that the national ranking is not reproduced at a single contributing site.

### Text-Encoder Embedding Classifiers

The three frozen-encoder classifiers were trained and evaluated under the same protocol as the six tabular families and are scored through the same code (Table 4). In the PUF test cohort none reached the highest AUROC or AUPRC for any outcome. Their shortfall against the best tabular value was 0.005 to 0.009 AUROC — 0.949 against 0.955 for mortality, 0.817 against 0.822 for any complication, 0.772 against 0.777 for unplanned return to OR, and 0.746 against 0.755 for readmission — and the confidence intervals did not overlap for mortality, any complication or readmission.

Measured against the individual families rather than the best of them, the picture is less uniform: the encoders were above logistic regression on all four outcomes and below the MLP and the FT-Transformer on all four, but the ordering against the remaining three families depended on the outcome. All three exceeded LightGBM for mortality (0.948–0.949 versus 0.946) and random forest for any complication, unplanned return to OR and readmission, and MedEmbed-large-v0.1 exceeded XGBoost for unplanned return to OR (0.772 versus 0.770); random forest in turn led all three for mortality. All three also exceeded the deployed ACS calculator for any complication (0.815–0.817 versus 0.802), with non-overlapping intervals, while the calculator led them for mortality (0.954).

Differences among the three encoders were smaller than their common distance from the tabular models, and an order of magnitude smaller than the range the tabular families span: 0.948–0.949 for mortality, 0.815–0.817 for any complication, 0.769–0.772 for unplanned return to OR and 0.745–0.746 for readmission. A 335M-parameter model trained specifically to produce embeddings and a 110M-parameter clinical masked-language model yielded similar discrimination on the same serialized record. In the single-institution cohort the intervals widened as they did for the tabular families, and one encoder row was best or tied-best on one metric: MedEmbed-large-v0.1 tied the MLP for readmission AUPRC (0.152), on 159 events.

Across the two cohorts and two metrics, no approach ranked first consistently. The FT-Transformer ranked first or tied for first on every AUROC comparison in the PUF cohort, but its margin over the MLP was 0.001 or less; under AUPRC the lead passed to LightGBM, XGBoost or the MLP depending on the outcome, and in the single-institution cohort the ordering dissolved into overlapping intervals. The apparent advantage of more complex architectures therefore depended on the outcome, the evaluation metric, and the evaluation cohort.

## Discussion

With the preoperative predictor set held constant, increasing model complexity produced only modest gains in discrimination in the national temporal test cohort. FT-Transformer achieved the highest or tied-highest AUROC across all four outcomes, but its advantage over logistic regression was small, ranging from 0.009 to 0.030. Differences among nonlinear models were smaller still; for example, XGBoost, LightGBM, MLP, and FT-Transformer achieved nearly identical AUROCs for any complication. Model ranking also depended on the evaluation metric: FT-Transformer led by AUROC, whereas LightGBM, XGBoost, and MLP achieved the highest AUPRC for different outcomes. In the single-institution cohort discrimination remained high, but confidence intervals widened and the national ordering was not reproduced. Together, these findings suggest that, once clinical information is fixed, architecture contributes less to predictive performance than might be inferred from comparisons in which both predictors and models change simultaneously; this analysis does not, however, speak to whether performance is equitable across sociodemographic subgroups (see Limitations).

Our finding that architecture contributed only modest gains once the predictor set was held constant is consistent with prior architecture-isolated comparisons on structured perioperative data. Shickel et al. found that deep neural networks offered incremental but not dramatic improvements over random forest and XGBoost for postoperative complications using a shared predictor set^6^, and Kim et al. similarly found that gradient-boosting and neural network approaches performed comparably to simpler models for postoperative pulmonary edema, with external validation showing preserved but attenuated discrimination^7^. Our results extend this pattern to a substantially larger national cohort and broader outcome set. Unlike Kim et al., however, we could not assess generalization to an independent institution: our single-institution cohort is drawn from a hospital that contributes to the PUF, so it does not provide the external check their design allowed.

The broader tabular-data literature disputes whether deep architectures reliably outperform tree-based ones, and our results do not settle it in either direction: they split by metric. Grinsztajn et al. found gradient-boosted trees competitive with or superior to deep architectures on medium-sized tabular datasets^10^. Here the deep models led on AUROC for all four outcomes, the FT-Transformer exceeding both XGBoost and LightGBM by 0.001 to 0.009. The trees led on AUPRC for two of them: LightGBM reached 0.302 for mortality against the FT-Transformer’s 0.286, and XGBoost and LightGBM reached 0.461 for any complication against 0.460. These margins are of the same order as the width of the confidence intervals, and in the single-institution cohort the ordering did not persist at all. On a cohort far larger than the medium-sized datasets Grinsztajn et al. examined, the tree-versus-deep question therefore has no stable answer here: which family leads depends on which metric is read.

In contrast, studies reporting larger performance gains from incorporating free-text clinical documentation — Lyu et al.’s multimodal transformer fusing clinical notes with structured EHR data^37^, and Chen et al.’s retrieval-augmented LLM approach for postoperative mortality^38^ — suggest that where substantial gains have been observed, they are more consistent with a modality effect (the addition of free-text information) than an architecture effect (the choice of model applied to the same structured inputs). Because our study held modality constant across all nine configurations, our results cannot speak to the magnitude of a modality effect directly, but the modest architecture effect we observed is consistent with the interpretation that the larger gains reported by Lyu et al. and Chen et al. likely reflect the additional information carried in free text rather than model complexity alone.

For the text-encoder embedding classifiers specifically, embeddings of the serialized record came close to the tabular models without matching them. That is the result reported by the two studies closest to this design. Gao et al. templated structured physiological values into text, extracted frozen embeddings and trained a conventional classifier on them, and found the raw numerical features still prevailed while the embeddings remained competitive^15^; Hegselmann et al. found plain-text EHR encodings embedded by modern encoders competitive with a dedicated EHR foundation model, yet with all training data available a gradient-boosting baseline on count features still edged both (macro-AUROC 0.777 versus 0.769) on the EHRSHOT benchmark^16^. Our predictor set is complete by construction — 69 curated, quality-audited registry fields with no free text — which is the regime in which both studies expect the tabular representation to hold its advantage, and it did. The result also sits between the two findings the broader literature offers: it is consistent with Jiang et al.’s result that an appropriately trained health system-scale language model can match task-specific models on clinical prediction tasks^17^, and it does not reproduce the large gap Brown et al. reported for zero-shot and few-shot GPT-3.5/GPT-4 prediction^18^ — training a classifier on frozen embeddings is a different and evidently stronger use of a language model than prompting one. But the shortfall, though small, was consistent: no encoder led on any outcome or metric in the national cohort.

The similarity of the three encoders is the more informative half of that result. Models differing threefold in parameter count and trained on different corpora, one of them specifically for embedding, yielded discrimination within 0.003 AUROC of each other on every outcome — a narrower spread than separates logistic regression from the FT-Transformer on the same cases. Within the three encoders evaluated, then, encoder choice had little effect on discrimination. That is a claim about these three checkpoints and not about clinical encoders in general: similar AUROC does not establish that the three representations encode the same information, and all three are 512-position BERT-family models, which forced the serialized record to be split into three passages and averaged. Recent work reports a clear margin between generations of encoder on plain-text EHR: Hegselmann et al. report Bio_ClinicalBERT at 0.705 macro-AUROC against 0.769 for a contemporary long-context embedding model on the same 15 tasks^16^. Whether a longer-context or more recent encoder would narrow the gap to the tabular models here is therefore untested and remains open.

The modest performance differences observed across tabular architectures have implications for perioperative model selection. When predictive gains are small, considerations beyond discrimination—including interpretability, computational requirements, implementation complexity, reproducibility, and ease of recalibration—may become increasingly important in selecting a model for deployment. The results also highlight the importance of holding the predictor set constant when benchmarking architectures. Apparent improvements attributed to a newer model may otherwise reflect differences in feature availability, preprocessing, or data modality rather than the modeling method itself. The lack of a stable architecture ranking across AUROC, AUPRC and the two evaluation cohorts further argues against selecting a model on a single performance metric evaluated in a single cohort, and against reading discrimination measured on a development sample as what a model will deliver at the sites where it runs — an independent evaluation of a widely deployed proprietary sepsis model found substantially worse discrimination than its developer had reported^39^.

### Limitations

This study has several limitations. PUF contains no patient-level identifiers, so prediction was necessarily performed at the operative-case level and repeated procedures by the same patient cannot be identified. The prespecified one-year gap increases the separation between the development and evaluation periods, but without a patient identifier it cannot establish patient-level independence, so residual patient-level overlap between the development and test cohorts cannot be excluded. Because PUF carries no institution identifier, heterogeneity in model performance across the contributing hospitals could not be assessed. Furthermore, variable definitions and availability changed across NSQIP years. Although annual files were harmonized and the prespecified predictor set was retained, several predictors were available only during part of the study period, introducing temporal differences in information availability. The study-defined complication composite retained occurrences flagged as present at the time of surgery and therefore differs from morbidity definitions that exclude such events. Additionally, this study did not evaluate model fairness or performance heterogeneity across sociodemographic subgroups, despite demographic variables (age, sex, race/ethnicity) being included as predictors; subgroup-level evaluation is an important direction for future work given known disparities in surgical outcomes and the documented capacity of clinical algorithms to encode them^5^. Finally, no independent external cohort was evaluated. The single-institution cohort is drawn from a hospital that participates in ACS NSQIP, and because the PUF is a systematic sample of participating sites, an unknown subset of those cases may also appear in the PUF evaluation year; since PUF carries no institution identifier, this overlap can be neither quantified nor removed. Those results therefore describe how the frozen models behave at one contributing site rather than how they generalize beyond the PUF sample, and the cohort is small — 3,490 cases with 23 deaths — so its confidence intervals are wide and cannot separate architectures. Generalizability to institutions not represented in the PUF sample remains untested, and performance measured on a national registry sample should not be assumed to transport to a given site^40^.

## Conclusion

In a national cohort of nearly 5 million surgical cases, increasing model complexity produced only modest gains in discrimination for perioperative outcome prediction once the preoperative predictor set was held fixed, and no single architecture — including the three pretrained clinical text-encoder embedding classifiers — consistently outperformed the others across outcomes or metrics. These findings suggest that, absent additional data modalities such as free-text documentation or imaging, the choice of tabular modeling architecture matters less than is often assumed, and that considerations such as interpretability, computational cost, and ease of recalibration may reasonably guide model selection when predictive differences are this small. Because every evaluation cohort in this benchmark is drawn from, or overlaps with, the national PUF sample, future work should assess whether these architecture rankings generalize to institutions outside it, alongside incorporating free-text clinical documentation and imaging within this same framework and testing longer-context encoders than the three 512-position checkpoints evaluated here.

## Data Availability

The ACS NSQIP Participant Use File used in this study is available to researchers upon request from the American College of Surgeons, subject to a Data Use Agreement. The Stony Brook University Hospital dataset used for single-institution evaluation is not publicly available due to institutional data-sharing restrictions and patient privacy considerations; requests for access may be directed to the corresponding author and are subject to Stony Brook University's institutional review and data use policies.

https://www.facs.org/quality-programs/data-and-registries/acs-nsqip/

