## Supplemental Table 1 for "From National Data to Local Evaluation: Benchmarking Machine Learning Models for Perioperative Risk Prediction"

**Supplementary Table S1. TRIPOD+AI Checklist**

*From National Data to Local Evaluation: Benchmarking Machine Learning Models for Perioperative Risk Prediction*

*Items marked with a footnote were not addressed in this study; these are disclosed as limitations in the manuscript where applicable.*

| **Section/Topic** | **Item** | **D/E** | **Checklist Item** | **Reported on Section/Paragraph** | **Notes** |
| --- | --- | --- | --- | --- | --- |
| **TITLE** | | | | | |
| Title | 1^[1]^ | D;E | Identify the study as developing or evaluating the performance of a multivariable prediction model, the target population, and the outcome to be predicted. | Title, page 1. "From National Data to Local Evaluation: Benchmarking Machine Learning Models for Perioperative Risk Prediction." | *"Local Evaluation" signals the single-institution comparison; target population and specific outcomes are not separately named in the title itself.* |
| **ABSTRACT** | | | | | |
| Abstract | 2 | D;E | See TRIPOD+AI for Abstracts checklist (completed separately below). | Abstract, page 1. | *See separate TRIPOD+AI for Abstracts checklist at the end of this document.* |
| **INTRODUCTION** | | | | | |
| Background | 3a | D;E | Explain the healthcare context (including whether diagnostic or prognostic) and rationale for developing or evaluating the prediction model, including references to existing models. | Introduction, paragraph 1 (ACS NSQIP Surgical Risk Calculator described as existing model; Bilimoria, Liu, Cohen cited). | *Reported, and strengthened with two additional calculator-validation citations (refs 2–3).* |
| Background | 3b^[2]^ | D;E | Describe the target population and the intended purpose of the prediction model in the context of the care pathway, including its intended users (e.g., healthcare professionals, patients, public). | Introduction, paragraph 1 (perioperative decision-making, informed consent, resource allocation). | *Partially reported — care-pathway context is described, but intended users are still not explicitly named.* |
| Background | 3c | D;E | Describe any known health inequalities between sociodemographic groups. | Introduction, paragraph 2 (Matar et al., Ann Surg Open, 2024, on sociodemographic disparities in ACS NSQIP outcomes; Obermeyer et al., Science, 2019, on algorithmic bias reproducing disparities). | *Cites two peer-reviewed sources and links the disparities discussion to the study’s own demographic predictors and to future fairness work.* |
| Objectives | 4 | D;E | Specify the study objectives, including whether the study describes the development or validation of a prediction model (or both). | Introduction, final paragraph ("The objective of this study is to compare the discriminative performance of traditional machine learning, conventional deep learning, and frozen clinical text-encoder embeddings..."). | *Reported.* |
| **METHODS** | | | | | |
| Data | 5a | D;E | Describe the sources of data separately for the development and evaluation datasets, the rationale for using these data, and representativeness of the data. | Methods, Study Design and Data Source and Cohort Definition (ACS NSQIP PUF, development/test split; single-institution cohort described with explicit non-independence caveat). | *Reported, and substantially strengthened — the single-institution cohort’s relationship to PUF (possible overlap) is now explicitly disclosed rather than presented as independent.* |
| Data | 5b | D;E | Specify the dates of the collected participant data, including start and end of participant accrual; and, if applicable, end of follow-up. | Methods, Cohort Definition (PUF 2018–2022 development, 2023 excluded, 2024 test; single-institution cohort 2024–2025). | *Reported.* |
| Participants | 6a^[3]^ | D;E | Specify key elements of the study setting (e.g., primary care, secondary care, general population) including the number and location of centres. | Methods, Study Design and Data Source ("more than 700 participating hospitals"); single-institution cohort described as "one academic medical center." | *Partially reported — hospital count and single-site status given, but care setting type (e.g., inpatient surgical) and geographic distribution are still not described.* |
| Participants | 6b | D;E | Describe the eligibility criteria for study participants. | Methods, Cohort Definition ("No additional eligibility criteria were applied, and no cases were excluded on clinical grounds."); harmonization exclusions for the single-institution cohort specified (2 cases). | *Reported.* |
| Participants | 6c^[4]^ | D;E | Give details of any treatments received, and how they were handled during model development or evaluation, if relevant. | NOT explicitly reported. | *Not addressed. Procedure/specialty is a predictor (Table 2), but treatment handling is not separately discussed.* |
| Data preparation | 7^[5]^ | D;E | Describe any data pre-processing and quality checking, including whether this was similar across relevant sociodemographic groups. | Methods, Predictors and Data Preprocessing (detailed, model-family-specific preprocessing; serialization and token-partitioning quality checks for the encoder arm). | *Preprocessing itself and its quality checks (e.g., "zero truncation was verified") are thoroughly described; consistency of preprocessing across sociodemographic groups specifically is still not addressed.* |
| Outcome | 8a^[6]^ | D;E | Clearly define the outcome being predicted and the time horizon, including how and when assessed, the rationale for choosing this outcome, and whether the method of outcome assessment is consistent across sociodemographic groups. | Methods, Outcomes (all four outcomes precisely defined with NSQIP variable names, 30-day windows, and full complication-composite variable list). | *Outcome definitions are exhaustively reported. Rationale for selecting these four specific outcomes and consistency of assessment across sociodemographic groups are still not addressed.* |
| Outcome | 8b^[7]^ | D;E | If outcome assessment requires subjective interpretation, describe the qualifications and demographic characteristics of the outcome assessors. | NOT reported. | *Not addressed. A brief reference to ACS Surgical Clinical Reviewer training/certification would satisfy this item.* |
| Outcome | 8c | D;E | Report any actions to blind assessment of the outcome to be predicted. | Methods, Outcomes ("Outcomes were recorded prospectively by NSQIP abstractors independent of this study’s model development, so no separate blinding procedure was required."). | *Reported.* |
| Predictors | 9a | D | Describe the choice of initial predictors (e.g., literature, previous models, all available predictors) and any pre-selection of predictors before model building. | Methods, Predictors and Data Preprocessing ("This predictor set was selected by a single clinician and co-author (A.R.K.) based on clinical relevance to perioperative risk, rather than derived from a previously published predictor set or risk model."). | *Reported.* |
| Predictors | 9b | D;E | Clearly define all predictors, including how and when they were measured (and any actions to blind assessment of predictors for the outcome and other predictors). | Methods, Predictors and Data Preprocessing; Table 2 (predictor domains); Table 3 (variable-level availability and harmonization, including the PRHEMOGLOBIN derivation formula). | *Reported, and strengthened — Table 3 now gives the exact derivation formula for the one cross-cohort measurement difference (PRHEMOGLOBIN), which is a strong, specific example of this item.* |
| Predictors | 9c^[8]^ | D;E | If predictor measurement requires subjective interpretation, describe the qualifications and demographic characteristics of the predictor assessors. | NOT reported. | *Not addressed; same consideration as item 8b.* |
| Sample size | 10 | D;E | Explain how the study size was arrived at (separately for development and evaluation), and justify that the study size was sufficient to answer the research question. Include details of any sample size calculation. | Methods, Cohort Definition ("No prospective sample size calculation was performed. Study size was instead determined by all eligible cases available in the ACS NSQIP PUF..."). | *Reported.* |
| Missing data | 11 | D;E | Describe how missing data were handled. Provide reasons for omitting any data. | Methods, Predictors and Data Preprocessing (sentinel value handling; per-model imputation; undefined-token handling for encoder serialization; missingness percentages reported in Table 1 footnote and Table 3). | *Reported, and strengthened with exact missingness figures (e.g., age missing for 1.21% of development cases).* |
| Analytical methods | 12a | D | Describe how the data were used (e.g., for development and evaluation of model performance) in the analysis, including whether the data were partitioned, considering any sample size requirements. | Methods, Cohort Definition and Models and Training (95%/5% fit/validation split; 2024 temporal test; single-institution cohort). | *Reported.* |
| Analytical methods | 12b | D | Depending on the type of model, describe how predictors were handled in the analyses (functional form, rescaling, transformation, or any standardisation). | Methods, Predictors and Data Preprocessing (model-family-specific handling, including the encoder serialization template and passage-partitioning procedure). | *Reported in detail.* |
| Analytical methods | 12c | D | Specify the type of model, rationale, all model building steps, including any hyperparameter tuning, and method for internal validation. | Methods, Models and Training and the new Model Parameters subsection (exact hyperparameter values given in full for all nine configurations — logistic regression, random forest, XGBoost, LightGBM, MLP, FT-Transformer, and the three encoder classifiers). | *Reported as a fully written-out Model Parameters subsection giving exact values for every model family, rather than as a table.* |
| Analytical methods | 12d | D;E | Describe if and how any heterogeneity in estimates of model parameter values and model performance was handled and quantified across clusters (e.g., hospitals, countries). | Discussion, Limitations ("Because PUF carries no institution identifier, heterogeneity in model performance across the contributing hospitals could not be assessed."). | *Explicitly stated.* |
| Analytical methods | 12e | D;E | Specify all measures and plots used (and their rationale) to evaluate model performance (e.g., discrimination, calibration, clinical utility) and, if relevant, to compare multiple models. | Methods, Statistical Analysis (AUROC, AUPRC with rationale for both given imbalance; bootstrap CIs); Figure 1 (AUROC forest-style plot across cohorts). | *Reported, and strengthened with Figure 1. Calibration explicitly not assessed, with rationale given.* |
| Analytical methods | 12f | E | Describe any model updating (e.g., recalibration) arising from the model evaluation. | Not applicable. | *Evaluation-only item; not applicable to this development-only study.* |
| Analytical methods | 12g | E | For model evaluation, describe how the model predictions were calculated (e.g., formula, code, object, API). | Not applicable; overlaps with item 22. | *Evaluation-only item; substantively addressed via the Model Parameters subsection and the GitHub code-availability statement (item 18f).* |
| Class imbalance | 13 | D;E | If class imbalance methods were used, state why and how this was done, and any subsequent methods to recalibrate the model or the model predictions. | Methods, Models and Training, class imbalance paragraph (cost-sensitive re-weighting fully described, including exact per-outcome weight ratios and an explicit citation on the harms of imbalance corrections for calibration). | *Reported, and strengthened with a supporting citation (van den Goorbergh et al.).* |
| Fairness | 14 | D;E | Describe any approaches that were used to address model fairness and their rationale. | Methods, Statistical Analysis ("No specific approach to address model fairness across sociodemographic subgroups... was undertaken in this benchmarking study."); Discussion, Limitations (expanded, with citation to Obermeyer et al. on algorithmic bias). | *This now explicitly states the approach taken (none) and situates that choice with a supporting citation, which satisfies the letter of this reporting item. Confirm with Zihan/Tahsin one final time whether any subgroup analysis was in fact run before submission, since no results are reported.* |
| Model output | 15 | D | Specify the output of the prediction model (e.g., probabilities, classification). Provide details and rationale for any classification and how the thresholds were identified. | Methods, Models and Training, class imbalance paragraph ("the resulting scores are not calibrated absolute risks... every operating point is rank-based rather than a fixed absolute-risk cut-off"). | *Reported.* |
| Training vs evaluation | 16 | D;E | Identify any differences between the development and evaluation data in healthcare setting, eligibility criteria, outcome, and predictors. | Methods, Predictors and Data Preprocessing; Table 3 (temporal and cross-cohort predictor differences, including the PRHEMOGLOBIN measurement difference); Table 1 (case-mix differences, e.g., sex distribution). | *Reported thoroughly — now the strongest-addressed item in the checklist, with both temporal (PUF-era) and cross-cohort (single-institution) differences explicitly tabulated.* |
| Ethical approval | 17 | D;E | Name the institutional research board or ethics committee that approved the study and describe the participant informed consent or the ethics committee waiver of informed consent. | Declarations block, Introduction (PUF: "its use did not require IRB review or approval"; single-institution cohort: "Use of unidentified Stony Brook University Hospital data was approved under a Stony Brook University Institutional Review Board exemption (IRB2026-00141)."). | *Both cohorts have an explicit ethical-approval statement.* |
| **OPEN SCIENCE** | | | | | |
| Funding | 18a | D;E | Give the source of funding and the role of the funders for the present study. | "This study received no external funding" (footnote-style block placed within the Introduction, before Methods, per AMIA proceedings formatting, which does not use a separate Declarations section). | *Reported.* |
| Conflicts of interest | 18b | D;E | Declare any conflicts of interest and financial disclosures for all authors. | "the authors declare no conflicts of interest" (same block as 18a). | *Reported.* |
| Protocol | 18c | D;E | Indicate where the study protocol can be accessed or state that a protocol was not prepared. | Represented via the OSF registration statement (item 18d); no separate protocol statement. | *Adequately reported via the registration DOI, consistent with earlier guidance that the OSF registration stands in for a separate protocol.* |
| Registration | 18d | D;E | Provide registration information for the study, including register name and registration number, or state that the study was not registered. | Methods, Study Design and Data Source ("prospectively registered on the Open Science Framework prior to model training and evaluation, Registration DOI: 10.17605/OSF.IO/JVKDH"). | *Reported.* |
| Data sharing | 18e | D;E | Provide details of the availability of the study data. | Declarations block, Introduction ("The ACS NSQIP PUF is available from the American College of Surgeons under a Data Use Agreement and is not redistributable by the authors. The single-institution dataset is not publicly available due to institutional data-sharing restrictions; requests may be directed to the corresponding author."). | *Covers both PUF and the single-institution dataset.* |
| Code sharing | 18f | D;E | Provide details of the availability of the analytical code. | "Analysis code is available at https://github.com/ZihanDing/Multi-Outcome-Perioperative-Risk-Prediction." (same block). | *A concrete public repository link is provided.* |
| **PATIENT AND PUBLIC INVOLVEMENT** | | | | | |
| Patient and public involvement | 19 | D;E | Provide details of any patient and public involvement during the design, conduct, reporting, interpretation, or dissemination of the study, or state no involvement. | "Patients and the public were not involved in this study." (same block). | *Reported.* |
| **RESULTS** | | | | | |
| Participants (Results) | 20a | D;E | Describe the flow of participants through the study, including the number of participants with and without the outcome and, if applicable, a summary of the follow-up time. A diagram may be helpful. | Table 1 (full outcome event counts and percentages for all four outcomes, across all three cohorts); Methods, Cohort Definition (cohort construction and exclusions narrated in text). | *Table 1 gives complete n (%) for every outcome in every cohort. No formal flow diagram is included; given the exhaustive tabular reporting and narrated exclusions, this is an optional enhancement rather than a gap.* |
| Participants (Results) | 20b | D;E | Report the characteristics overall and, where applicable, for each data source or setting, including key dates, key predictors (including demographics), treatments received, sample size, number of outcome events, follow-up time, and amount of missing data. A table may be helpful. | Table 1 (age mean/SD/median, sex distribution including non-binary/intersex categories, outcome frequencies, and missingness, for all three cohorts). | *Fully addressed with a baseline characteristics table.* |
| Participants (Results) | 20c | E | For model evaluation, show a comparison with the development data of the distribution of important predictors (demographics, predictors, and outcome). | Table 1 (case-mix comparison across development, temporal test, and single-institution cohorts, e.g., 69.2% vs. 57.9% female). | *Now substantively addressed even though formally an evaluation-only item, since the single-institution cohort’s distribution is directly compared to PUF’s in Table 1.* |
| Model development | 21 | D;E | Specify the number of participants and outcome events in each analysis (e.g., for model development, hyperparameter tuning, model evaluation). | Table 1 (exact n and % for every outcome, in the development, temporal test, and single-institution cohorts). | *Reported.* |
| Model specification | 22 | D | Provide details of the full prediction model (e.g., formula, code, object, API) to allow predictions in new individuals and to enable third party evaluation and implementation, including any restrictions to access or reuse. | Methods, Model Parameters subsection (full hyperparameter values for all nine configurations); Declarations, Code Availability (public GitHub repository link). | *Both the parameter values and a public code repository are provided, which together substantively satisfy this item.* |
| Model performance | 23a^[9]^ | D;E | Report model performance estimates with confidence intervals, including for any key subgroups (e.g., sociodemographic). Consider plots to aid presentation. | Table 4 (AUROC/AUPRC with 95% CIs for all nine models, both cohorts); Figure 1 (visual forest-style comparison); ACS NSQIP calculator included as a deployed-tool reference. | *Overall performance reporting is now excellent — comprehensive, well-visualized, and benchmarked against the deployed clinical tool. No subgroup (sociodemographic) performance breakdown is included, consistent with the fairness item (14).* |
| Model performance | 23b | D;E | If examined, report results of any heterogeneity in model performance across clusters. | Results, Single-Institution Cohort subsection functions as a de facto single-cluster heterogeneity check, though not framed in TRIPOD-Cluster terms. | *Substantively addressed by the single-institution comparison, even though PUF itself has no institution-level cluster analysis.* |
| Model updating | 24 | E | Report the results from any model updating, including the updated model and subsequent performance. | Not applicable. | *Evaluation-only item; not applicable to this development-only study.* |
| **DISCUSSION** | | | | | |
| Interpretation | 25 | D;E | Give an overall interpretation of the main results, including issues of fairness in the context of the objectives and previous studies. | Discussion, paragraph 1 (Principal Findings), final clause ("this analysis does not, however, speak to whether performance is equitable across sociodemographic subgroups (see Limitations)."); extensive literature-comparison paragraphs follow. | *The fairness caveat is cross-referenced directly within the main interpretive paragraph, not confined only to Limitations.* |
| Limitations | 26 | D;E | Discuss any limitations of the study (such as a non-representative sample, sample size, overfitting, missing data) and their effects on any biases, statistical uncertainty, and generalisability. | Discussion, Limitations (patient-level identification, temporal variable availability, complication composite definition, fairness/subgroup evaluation with citation, single-institution cohort non-independence, wide CIs, generalizability caveat with citation). | *Each limitation is tied to a specific consequence rather than stated abstractly.* |
| Usability | 27a^[10]^ | D | Describe how poor quality or unavailable input data (e.g., predictor values) should be assessed and handled when implementing the prediction model. | Methods, Predictors and Data Preprocessing describes the imputation/undefined-token approach used during model development and evaluation. | *Partially reported — the development-time handling of missing/poor-quality data is thoroughly described, but explicit guidance for a deployment setting (i.e., what a future implementer should do) is not framed separately.* |
| Usability | 27b^[11]^ | D | Specify whether users will be required to interact in the handling of the input data or use of the model, and what level of expertise is required of users. | NOT reported. | *Missing. No discussion of user interaction requirements or expertise needed to operate the model in practice.* |
| Usability | 27c | D;E | Discuss any next steps for future research, with a specific view to applicability and generalisability of the model. | Conclusion (future work: external validation at institutions outside the PUF sample, incorporating free-text documentation and imaging, testing longer-context encoders). | *Reported, and strengthened with a specific, concrete future-work item (longer-context encoders) grounded in the Discussion’s Hegselmann et al. citation.* |

**TRIPOD+AI for Abstracts**

| **Item** | **Checklist Item** | **Reported** | **Notes** |
| --- | --- | --- | --- |
| 1^[12]^ | Identify the study as developing or evaluating the performance of a multivariable prediction model, the target population, and the outcome to be predicted. | Abstract, sentence 1. | *Reported at a general level; specific target population (surgical patients) still not named explicitly.* |
| 2 | Provide a brief explanation of the healthcare context and rationale for developing or evaluating the performance of all models. | Abstract, sentence 1 ("Whether newer modeling approaches improve perioperative risk prediction is unclear."). | *Reported.* |
| 3 | Specify the study objectives, including whether the study describes model development, evaluation, or both. | Abstract, sentence 2 ("we benchmarked six tabular model families and three pretrained clinical text-encoder embedding classifiers..."). | *Reported.* |
| 4 | Describe the sources of data. | Abstract, sentence 2 (4,995,670 ACS NSQIP cases 2018–2022 training, 963,565 cases 2024 test). | *Reported.* |
| 5^[13]^ | Describe the eligibility criteria and setting where the data were collected. | NOT explicitly in Abstract. | *Setting (multi-institutional national registry) is not stated in the Abstract itself, though implied by "ACS NSQIP."* |
| 6 | Specify the outcome to be predicted by the model, including time horizon of predictions in case of prognostic models. | Abstract, sentence 2 ("four 30-day outcomes"). | *Reported.* |
| 7^[14]^ | Specify the type of model, a summary of the model-building steps, and the method for internal validation. | Abstract, sentence 2 (six tabular model families and three encoder classifiers named). | *Model types are named; internal validation method (early stopping/validation split) is not mentioned in the Abstract.* |
| 8 | Specify the measures used to assess model performance (e.g., discrimination, calibration, clinical utility). | Abstract (AUROC reported throughout; AUPRC referenced via "rankings changed under AUPRC"). | *Reported.* |
| 9^[15]^ | Report the number of participants and outcome events. | Abstract, sentence 2 (4,995,670 development / 963,565 test cases; 3,490-case single-institution check). | *Cohort sizes are reported; outcome event counts are not given in the Abstract itself.* |
| 10 | Summarise the predictors in the final model. | Abstract, sentence 1 ("73 prespecified preoperative variables — 69 observed and used as tabular inputs, all 73 serialized to text"). | *Precisely quantified, including the 69-vs-73 accounting.* |
| 11^[16]^ | Report model performance estimates (with confidence intervals). | Abstract (AUROC ranges and point differences reported, e.g., "0.755–0.955," "exceeding logistic regression by only 0.009–0.030"); confidence intervals not given in the Abstract itself. | *Point estimates and ranges are reported in more precise detail than before; individual confidence intervals still appear only in Table 4 in the main text, not the Abstract.* |
| 12 | Give an overall interpretation of the main results. | Abstract, final sentence ("Once the predictor set is fixed, architecture and encoder choice move discrimination little."). | *The Abstract ends with a clear interpretive takeaway sentence.* |
| 13^[17]^ | Give the registration number and name of the registry or repository. | NOT in Abstract text; registration DOI appears in Methods and in the footnote-style Declarations block, not within the Abstract itself. | *Confirm AMIA formatting requirements — some venues want this in the abstract block itself rather than elsewhere in the manuscript.* |

1. Not addressed in this study.
2. Not addressed in this study.
3. Not addressed in this study.
4. Not addressed in this study.
5. Not addressed in this study.
6. Not addressed in this study.
7. Not addressed in this study.
8. Not addressed in this study.
9. Not addressed in this study.
10. Not addressed in this study.
11. Not addressed in this study.
12. Not addressed in this study.
13. Not addressed in this study.
14. Not addressed in this study.
15. Not addressed in this study.
16. Not addressed in this study.
17. Not addressed in this study.
